# Assessing performance and clinical usefulness in prediction models with survival outcomes: practical guidance for Cox proportional hazards models

**DOI:** 10.1101/2022.03.17.22272411

**Authors:** David J McLernon, Daniele Giardiello, Ben Van Calster, Laure Wynants, Nan van Geloven, Maarten van Smeden, Terry Therneau, Ewout W Steyerberg, topic groups 6 and 8 of the STRATOS Initiative

## Abstract

Risk prediction models need thorough validation to assess their performance. Validation of models for survival outcomes poses challenges due to the censoring of observations and the varying time horizon at which predictions can be made. We aim to give a description of measures to evaluate predictions and the potential improvement in decision making from survival models based on Cox proportional hazards regression.

As a motivating case study, we consider the prediction of the composite outcome of recurrence and death (the ‘event’) in breast cancer patients following surgery. We develop a Cox regression model with three predictors as in the Nottingham Prognostic Index in 2982 women (1275 events within 5 years of follow-up) and externally validate this model in 686 women (285 events within 5 years). The improvement in performance was assessed following the addition of circulating progesterone as a prognostic biomarker.

The model predictions can be evaluated across the full range of observed follow up times or for the event occurring by a fixed time horizon of interest. We first discuss recommended statistical measures that evaluate model performance in terms of discrimination, calibration, or overall performance. Further, we evaluate the potential clinical utility of the model to support clinical decision making. SAS and R code is provided to illustrate apparent, internal, and external validation, both for the three predictor model and when adding progesterone.

We recommend the proposed set of performance measures for transparent reporting of the validity of predictions from survival models.

## Introduction

Prediction models for survival outcomes are important for clinicians who wish to estimate a patient’s risk (i.e. probability) of experiencing a future outcome. The term ‘survival’ outcome is used to indicate any prognostic or time-to-event outcome, such as death, progression, or recurrence of disease. Such risk estimates for future events can support shared decision making for interventions in high-risk patients, help manage the expectations of patients, or stratify patients by disease severity for inclusion in trials.^1^ For example, a prediction model for persistent pain after breast cancer surgery might be used to identify high risk patients for intervention studies.^2^

Once a prediction model has been developed it is common to first assess its performance for the underlying population. This is referred to as internal validation, which can be performed using the dataset on which the model was developed, for example by cross-validation or bootstrapping techniques.^3^ External validation refers to performance in a plausibly related population, which requires an independent dataset which may differ in setting, time or place.^4, 5^

Ample guidance exists for assessing the performance of prediction models for binary outcomes, where the logistic regression model is most commonly used for model development.^6–8^ Validation of a survival model poses more of a challenge due to the censoring of observation times when a patient’s outcome is undetermined during the study period. If assessing 5-year survival, for instance, some subjects may have less than 5 years of follow-up without experiencing the event of interest.^3^ Moreover, predictions can be evaluated over the entire range of observed follow up times or for the event occurring by a fixed time horizon of interest. The international STRengthening Analytical Thinking for Observational Studies (STRATOS) initiative (http://stratos-initiative.org) began in 2013 and aims to provide accessible and accurate guidance documents for relevant topics in the design and analysis of observational studies.^9^

This STRATOS article aims to provide guidance for assessing discrimination, calibration, and clinical usefulness for survival models, building on the methodological literature for survival model evaluation.^10–12^ For illustration, we consider the performance of a Cox model to predict recurrence free survival (i.e. being alive and without breast cancer recurrence) at 5 years in breast cancer patients. We also describe how to assess the improvement in predictive ability and decision-making when adding a prognostic biomarker.

## Methods and Results

In the following, we discuss three key issues for the evaluation of predictions from survival prediction models. We then describe our breast cancer case study, present how we can predict survival outcomes with the Cox proportional hazards model, perform validation of predictions, and assess the potential clinical usefulness of a prediction model.

### Key issues when validating a survival model

Three major issues differentiate the validation of survival models from models for binary outcomes. First, we need to decide on a time point or time range over which to assess the validation. This choice needs to be grounded in both the available data and the intended practical use of the model predictions. Altman considers a case where a model will be used for individual risk stratification in advanced pancreatic cancer patients.^13^ In such a case a quite short time horizon is indicated of e.g. 18 months. Other situations with longer follow-up might use 3, 5, 10, or even 20 years.

A second issue is whether to consider prediction only up to *a fixed time point* or over an entire range of follow-up. In our case study we focus on 5 years from enrollment as the upper limit. For a cutoff of 5 years, we need to decide if only the binary outcome of whether the event occurred before or after 5 years is of interest, or also the ability to distinguish between survival of 1 and 4 years, for instance. We will give measures of performance for both settings.

A last technical issue is that estimation of the baseline survival S_0_(*t*) from the Cox model is necessary for full validation of a prediction model. However, many published reports do not provide this function (see Box 1 for further details).^10^

#### Box 1 The Cox proportional hazards model to make predictions for new patients

Hazard ratios express how baseline patient characteristics (or predictors) are associated with the hazard rate, that is the instantaneous rate of the event occurring at time *t*, having survived until time *t*. Mathematically, the Cox model for the hazard rate, h(t), is

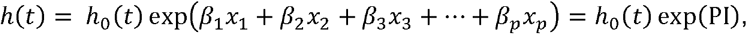

where the *β*’s are regression coefficients for the *p* predictors *x*_*1*_ to *x*_*p*_ (e.g., the patient’s age, disease stage, comorbidity). These regression coefficients are the log of the hazard ratios. The prognostic index, PI, represents the sum of the regression coefficients multiplied by the value of their respective predictors. The Cox model assumes that hazards for different values of a predictor are proportional during follow-up. For example, if the hazard of the event for patient A is half that of patient B at time *t*, the hazard ratio of 0.5 holds for these two patients at any other time point.

The baseline hazard function *h*_0_(*t*) is the same for all patients analogous to the intercept in linear or logistic regression models. If the primary focus of an analysis is relative risk estimation, the Cox model can be used to obtain hazard ratios without worrying about baseline hazard estimation. For estimating the risk that a patient experiences the event, i.e. absolute risk estimation, we require the baseline survival function *S*_*0*_*(t)* which is the predicted risk of survival for the patient whose predictor values are the reference categories (for categorical predictors) or zero/the mean (for continuous predictors). By integrating the hazard function from time 0 to *t* we obtain the cumulative hazard function, *H*(*t*) = *H*_0_(*t*)exp(*PI*), where *H*_0_(*t*) is the baseline cumulative hazard function. *H*_0_(*t*) is then used to estimate the probability of survival up to time *t*, i.e. not experiencing the event up to time t:

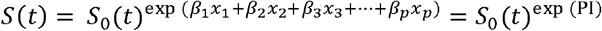

where *S*_0_(*t*) = exp(−*H*_0_(*t*)), the baseline survival at time *t* (e.g., *t* = 5 years after surgery). The absolute risk of an event within *t* years is calculated as 1 – *S(t)*. The baseline hazard of a Cox model is often estimated non-parametrically in contrast to parametric survival models such as the accelerated failure time model.

Estimates of absolute risk are necessary for many of the performance measures discussed below. A model development study hence needs to have reported the baseline hazard function or baseline survival function, or at least survival at the time point of interest, and a specification of calculation of the PI. This is analogous to a logistic regression model to predict a binary outcome, which additionally needs reporting of a model intercept rather than only odds ratios.

### Description of the case study

We analysed data from patients who had primary surgery for breast cancer between 1978 and 1993 in Rotterdam.^14, 15^ Patients were followed until 2007. After exclusions, 2982 patients were included in the model development cohort (Table 1). The outcome was recurrence-free survival, defined as time from primary surgery to recurrence or death. Over the maximum follow-up time of 19.3 years, 1,713 events occurred, and the estimated median potential follow-up time, calculated using the reverse Kaplan-Meier method, was 9.3 years.^16^ Out of 2,982 patients, 1,275 suffered a recurrence or death within the follow-up time of interest, which was 5 years, and 126 were censored before 5 years. An external validation cohort consisted of 686 patients with primary node positive breast cancer from the German Breast Cancer Study Group,^17^ where 285 suffered a recurrence or died within 5 years of follow-up, and 280 were censored before 5 years. Five year predictions were chosen as that was the lowest median survival from the two cohorts (Rotterdam cohort, 6.7 years; German cohort, 4.9 years).

**Table 1.** Characteristics of the breast cancer cohorts used for model development and external validation^14, 17^.

| Characteristic |  | Development cohort<br>(n=2982, 1275<br>events <5 years) | Validation cohort<br>(n=686, 285 events<br><5 years) |
| --- | --- | --- | --- |
| Size (mm) | ≤20 | 1387 (46.5) | 180 (26.2) |
|  | 21-50 | 1291 (43.3) | 453 (66.0) |
|  | >50 | 304 (10.2) | 53 (7.7) |
| Number of Nodes | 0 | 1436 (48.2) | 0 (0.0) |
|  | 1 to 3 | 764 (25.6) | 376 (54.8) |
|  | >3 | 782 (26.2) | 310 (45.2) |
| Grade of Tumour | 1 or 2 | 794 (26.6) | 525 (76.5) |
|  | 3 | 2188 (73.4) | 161 (23.5) |
| Age (years: median (IQR)) |  | 54 (45 to 65) | 53 (46 to 61) |
| Circulating progesterone (PGR,<br>ng/mL: median (IQR)) |  | 41 (4 to 198) | 33 (7 to 132) |

Numbers (%) unless otherwise stated

### Prediction of survival outcomes

The Cox proportional hazards model is a standard for analysing survival data in biomedical settings^18^ A Cox model estimates log hazard ratios, but for prediction, estimation of the baseline survival is also required. Both are needed for a full assessment of performance of a survival model in new patients (external validation, Box 1).

### Model development in the case study

A Cox regression model was fit to estimate recurrence free survival using three predictors: number of lymph nodes (0, 1-3, >3), tumour size (≤20mm, 21-50mm, >50mm) and pathological grade (1, 2, 3, see Table 2). Although we emphasize that it is generally poor practice to categorise continuous variables, tumour size was not available in continuous form in the dataset, and number of lymph nodes was categorised to match its form in the well-known Nottingham Prognostic Index.^19,20^ Since we were interested in predictons up to 5 years, we applied administrative censoring at 5 years. The Cox model assumes that hazards for different values of a predictor are proportional during follow-up. While found some evidence of non-proportional hazards (p<0.001, Grambsch and Therneau global test), we chose to ignore this violation here since it was relatively minor at graphical inspection. Furthermore, predictions made at the time of administrative censoring (5 years here) have been shown to be robust regardless of such violations.^21^

**Table 2.** Cox regression models predicting event free survival in Rotterdam breast cancer development dataset (n=2982), without and with PGR.

|  | Without PGR | With PGR |
| --- | --- | --- |
|  | Hazard ratio<br>(95% CI) | Hazard ratio<br>(95% CI) |
| Size (mm) |  |  |
| ≤20 | 1 | 1 |
| 21-50 | 1.47 (1.29 to 1.67) | 1.44 (1.26 to 1.63) |
| >50 | 1.94 (1.62 to 2.32) | 1.90 (1.59 to 2.27) |
| Number of nodes |  |  |
| 0 | 1 | 1 |
| 1 to 3 | 1.43 (1.24 to 1.66) | 1.46 (1.26 to 1.70) |
| >3 | 2.89 (2.52 to 3.32) | 2.88 (2.51 to 3.31) |
| Tumour grade |  |  |
| 1 or 2 | 1 | 1 |
| 3 | 1.46 (1.27 to 1.67) | 1.37 (1.19 to 1.58) |
| PGR (ng/ml) |  | 1.46<sup>§</sup> (1.27 to 1.68) |
| PGR1<sup>§</sup> |  |  |

**For model without PGR, the formula for the prognostic index is:**
The survival at 5 years can be calculated as:

The formula for the prognostic index was estimated as follows:

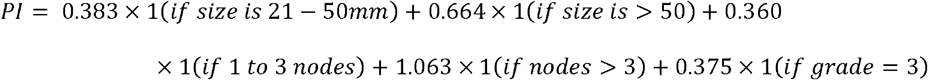

The probability of experiencing the event within 5 years can be calculated as:

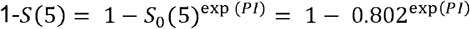

The baseline survival at 5 years (0.802) applies to the reference categories for the three predictors in the model (see R and SAS code in https://github.com/danielegiardiello/Prediction_performance_survival). So, a woman with a tumor size <=20mm, no nodes, and grade<3, has an estimated risk of 1 – 0.802^1^ = 19.8% of recurrence or breast cancer mortality within 5 years.

### Measures of performance

Model performance was assessed in the development dataset (apparent validation) and in the German dataset (external validation). Internal validation was assessed using the bootstrap resampling approach which provides stable estimates of performance for the population where the sample originated from. The difference between the apparent performance and the internal performance represents the “optimism” in performance of the original model (see Appendix 1 for further details).

### Discrimination

A first question is how well the model predictions separate high from low risk patients: discriminative ability. Patients with an earlier event time should exhibit a higher risk and those with later event time a lower risk.

#### Fixed time point discrimination

Measures that assess the prediction by a fixed time point are the similar to those for binomial outcomes. A primary issue that arises, however, is censoring in the validation data set. If we choose an evaluation time of 5 years, for instance, how are subjects who are censored before 5 years in the validation set to be assessed? For these we have a predicted risk at 5 years from the model, but do not have an observed value of the outcome at 5 years. One approach is to use inverse probability of censoring weights (IPCW), to reassign the case weights of those censored to other observations with longer follow up (see Table S1).

Uno applies such inverse weights, and this is our recommended method for assessing discrimination at a fixed time point, though many others exist.^22, 23^ It assesses all pairs of patients where one experiences the event before the chosen time point and the other remains event free up to that time and calculates the proportion of those pairs for which the first mentioned patient has highest estimated risk (Table S2). Uno’s IPCW approach for 5 year prediction was 0.71 [95% CI 0.69 to 0.73] at model development (apparent validation). Internal validation suggested no statistical optimism (remained 0.71 using 500 bootstrap samples), while external validation showed a slightly poorer performance (0.69 [95% CI 0.63 to 0.75], Table 3).

**Table 3.**
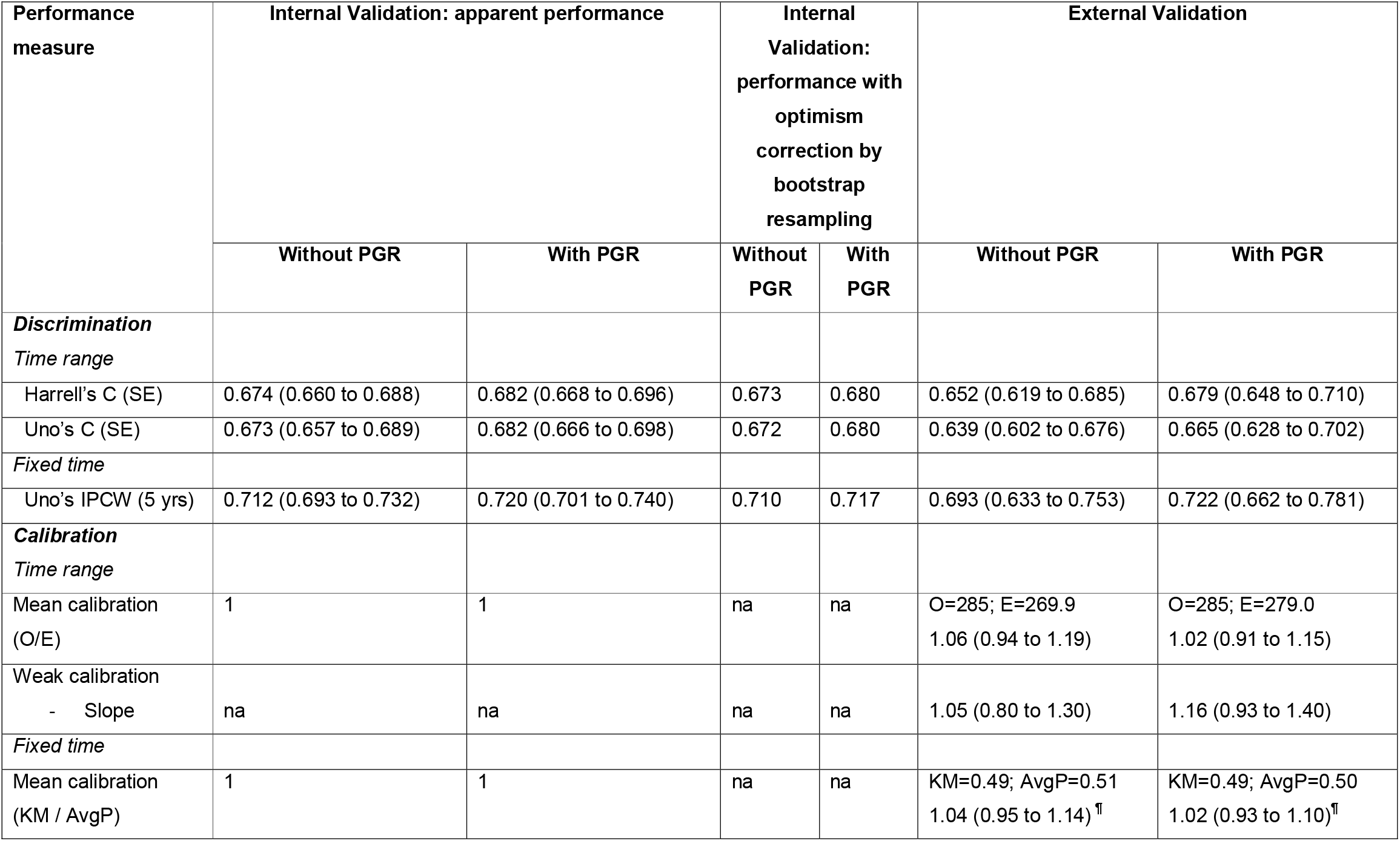
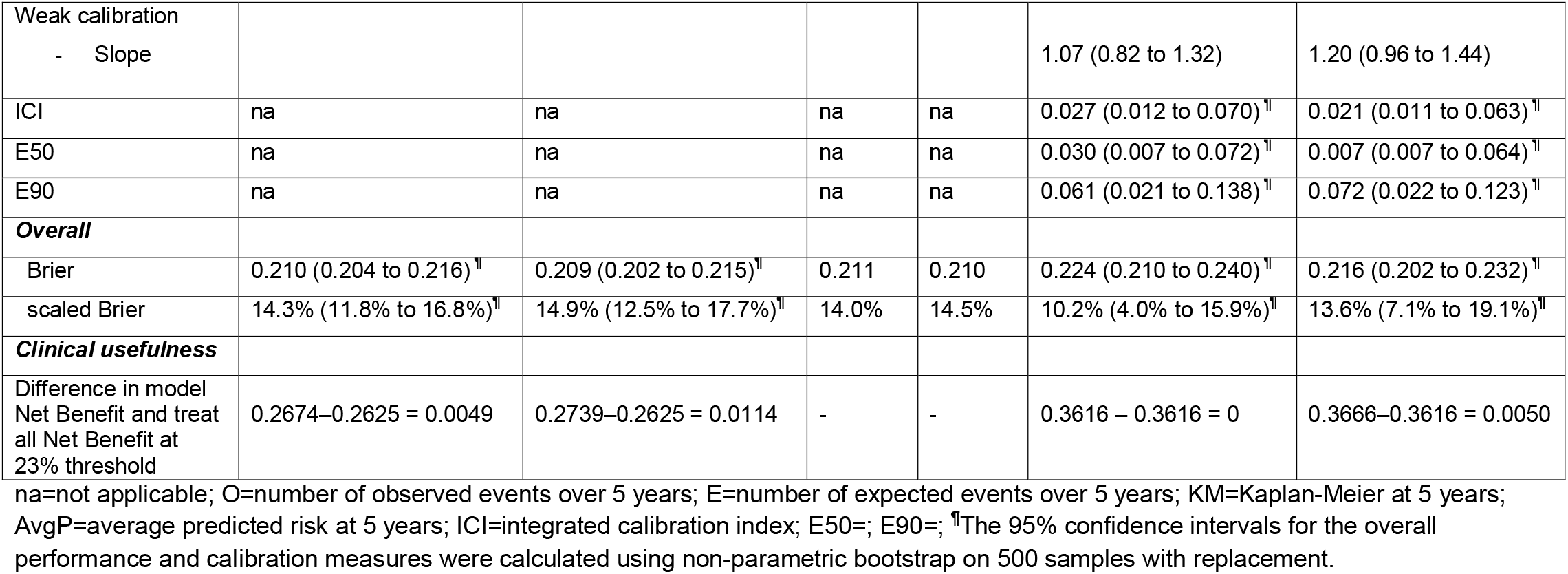
Performance of breast cancer model with and without PGR at 5 years in development (n=2982) and validation data (n=686)

| Performance measure | Internal Validation: apparent performance |  | Internal Validation: performance with optimism correction by bootstrap resampling |  | External Validation |  |
| --- | --- | --- | --- | --- | --- | --- |
|  | Without PGR | With PGR | Without PGR | With PGR | Without PGR | With PGR |
| **Discrimination** |  |  |  |  |  |  |
| *Time range* |  |  |  |  |  |  |
| Harrell's C (SE) | 0.674 (0.660 to 0.688) | 0.682 (0.668 to 0.696) | 0.673 | 0.680 | 0.652 (0.619 to 0.685) | 0.679 (0.648 to 0.710) |
| Uno's C (SE) | 0.673 (0.657 to 0.689) | 0.682 (0.666 to 0.698) | 0.672 | 0.680 | 0.639 (0.602 to 0.676) | 0.665 (0.628 to 0.702) |
| *Fixed time* |  |  |  |  |  |  |
| Uno's IPCW (5 yrs) | 0.712 (0.693 to 0.732) | 0.720 (0.701 to 0.740) | 0.710 | 0.717 | 0.693 (0.633 to 0.753) | 0.722 (0.662 to 0.781) |
| **Calibration** |  |  |  |  |  |  |
| *Time range* |  |  |  |  |  |  |
| Mean calibration (O/E) | 1 | 1 | na | na | O=285; E=269.9<br>1.06 (0.94 to 1.19) | O=285; E=279.0<br>1.02 (0.91 to 1.15) |
| Weak calibration - Slope | na | na | na | na | 1.05 (0.80 to 1.30) | 1.16 (0.93 to 1.40) |
| *Fixed time* |  |  |  |  |  |  |
| Mean calibration (KM / AvgP) | 1 | 1 | na | na | KM=0.49; AvgP=0.51<br>1.04 (0.95 to 1.14)<sup>¶</sup> | KM=0.49; AvgP=0.50<br>1.02 (0.93 to 1.10)<sup>¶</sup> |
| Weak calibration<br>- Slope |  |  |  |  | 1.07 (0.82 to 1.32) | 1.20 (0.96 to 1.44) |
| ICI | na | na | na | na | 0.027 (0.012 to 0.070)<sup>¶</sup> | 0.021 (0.011 to 0.063)<sup>¶</sup> |
| E50 | na | na | na | na | 0.030 (0.007 to 0.072)<sup>¶</sup> | 0.007 (0.007 to 0.064)<sup>¶</sup> |
| E90 | na | na | na | na | 0.061 (0.021 to 0.138)<sup>¶</sup> | 0.072 (0.022 to 0.123)<sup>¶</sup> |
| **Overall** |  |  |  |  |  |  |
| Brier | 0.210 (0.204 to 0.216)<sup>¶</sup> | 0.209 (0.202 to 0.215)<sup>¶</sup> | 0.211 | 0.210 | 0.224 (0.210 to 0.240)<sup>¶</sup> | 0.216 (0.202 to 0.232)<sup>¶</sup> |
| scaled Brier | 14.3% (11.8% to 16.8%)<sup>¶</sup> | 14.9% (12.5% to 17.7%)<sup>¶</sup> | 14.0% | 14.5% | 10.2% (4.0% to 15.9%)<sup>¶</sup> | 13.6% (7.1% to 19.1%)<sup>¶</sup> |
| **Clinical usefulness** |  |  |  |  |  |  |
| Difference in model<br>Net Benefit and treat<br>all Net Benefit at<br>23% threshold | 0.2674–0.2625 = 0.0049 | 0.2739–0.2625 = 0.0114 | - | - | 0.3616 – 0.3616 = 0 | 0.3666–0.3616 = 0.0050 |

na=not applicable; O=number of observed events over 5 years; E=number of expected events over 5 years; KM=Kaplan-Meier at 5 years; AvgP=average predicted risk at 5 years; ICI=integrated calibration index; E50=; E90=; <sup>¶</sup>The 95% confidence intervals for the overall performance and calibration measures were calculated using non-parametric bootstrap on 500 samples with replacement.

#### Time range discrimination

Harrell’s concordance index (C) is commonly used to assess global performance.^24^ It is calculated as a fraction where the denominator is the number of all possible pairs of patients in which one patient experiences the event first and the other later. Harrell’s C quantifies the degree of concordance as the proportion of such pairs where the patient with a longer survival time has better predicted survival (lower PI). Using our time range of 0 to 5 years, Harrell’s C was 0.67 [95% CI 0.66 to 0.69] at apparent validation. Again, no optimism was noted (C=0.67) and a slightly lower performance at external validation (C=0.65 [95% CI 0.62 to 0.69]). Uno’s C uses a time dependent weighting that more fully adjusts for censoring (more details in appendix 2).^25^ Uno’s C was also 0.67 [95% CI 0.66 to 0.69] at apparent validation, 0.67 at internal validation and 0.64 [95% CI 0.60 to 0.68] for external validation in our case study.

### Calibration

A second important question to answer when validating a model is ‘how well do observed outcomes agree with model predictions? This relates to calibration.^8, 11^ Assessment of calibration is essential at external validation ^3, 26^. Below we describe a hierarchy of calibration levels and its application to survival model predictions, in line with a previously proposed framework.^8^

#### Mean calibration

Mean calibration (or calibration-in-the-large) refers to agreement of the predicted and observed survival fraction.

Fixed time point mean calibration is typically expressed in terms of the ratio of the observed survival fraction and the average predicted risk. The observed survival fraction at the chosen time point needs to be estimated due to censoring, which can be done using the Kaplan-Meier estimator. For the external validation cohort, the Kaplan-Meier estimate of experiencing the event within 5 years was 51%, while the average predicted probability was 49%. This indicates a minor deviation from perfect mean calibration (a ratio of 1.04, 95% CI [0.95 to 1.14], Table 3).

#### Weak calibration

The term ‘weak’ refers to the limited flexibility in assessing calibration. We are essentially summarising calibration of the observed proportions of outcomes versus predicted probabilities using only two parameters i.e. a straight line. In other words, perfect weak calibration is defined as mean calibration ratio and calibration slope of unity. Mean calibration indicates systematic underprediction or overprediction. The calibration slope indicates the overall strength of the PI, which can be interpreted as the level of overfitting (slope <1) or underfitting (slope>1).

For a fixed time point assessment of weak calibration, we can predict the outcome at 5 years for every patient but we need to determine the observed outcome at 5 years even for those who were censored before that time. One way to do this is to fit a new ‘secondary’ Cox model using all of the validation data with the PI from the development model as the only covariate. The calibration slope is the coefficient of the PI. In our case study it was 1.07 [95% CI 0.82 to 1.32] for the 5 year predictions, confirming very good calibration.

#### Moderate calibration

Moderate calibration concerns whether among patients with the same predicted risk, the observed event rate equals the predicted risk.^6^ A smooth calibration curve of the observed event rates against the predicted risks is used for assessment of moderate calibration.

The relation between the outcome at a fixed time point and predictions can be visualised by plotting the predicted risk from another ‘secondary’ Cox model against the predicted risk from the development model.^27^ The details are presented in Appendix 3 and Table S1.

The calibration plot shows good agreement between predictions from the developed model and observed event rates as estimated by the secondary model (Fig 1A). This plot can be characterized further by some calibration metrics. The Integrated Calibration Index (ICI) is the mean absolute difference between smoothed observed proportions and predicted probabilities. The E50 and E90 denote the median and the 90th percentile absolute difference between observed and predicted probabilities of the outcome.^27^ For our validation cohort, we estimated ICI was 0.03 [95% CI 0.01 to 0.07], E50=0.03 [95% CI 0.007 to 0.07] and E90=0.06 [95% CI 0.02 to 0.14].

**Figure 1.**
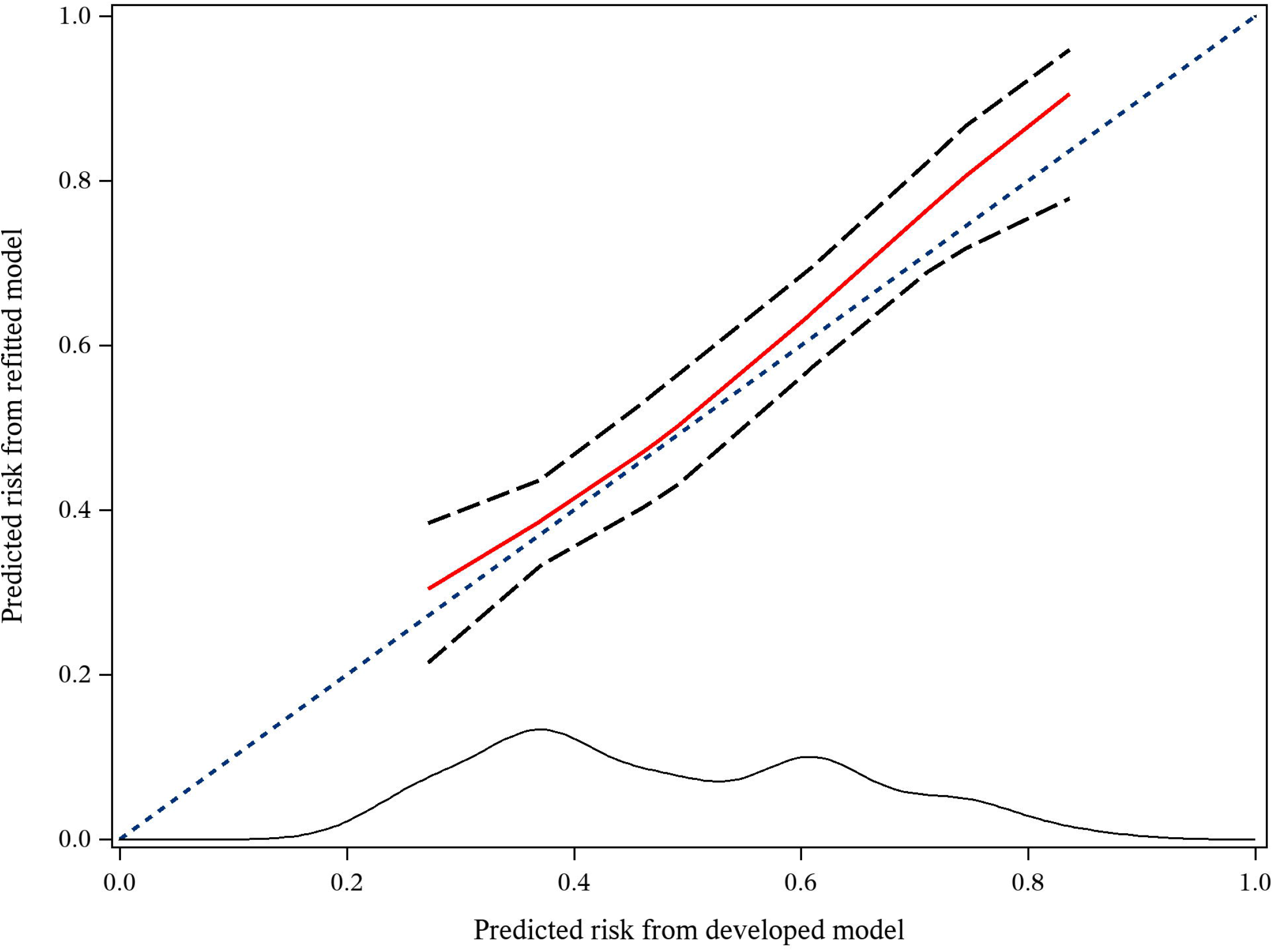
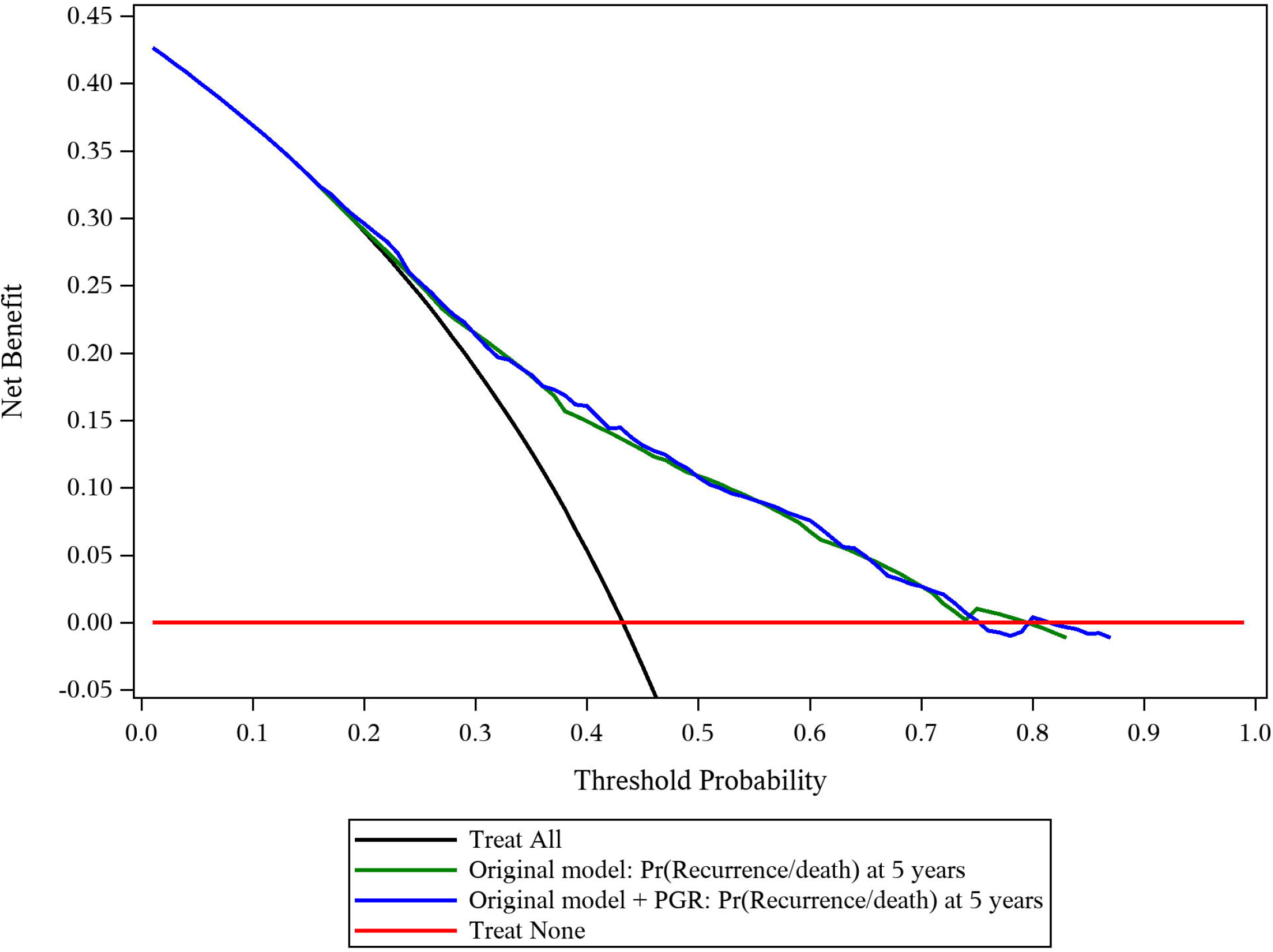
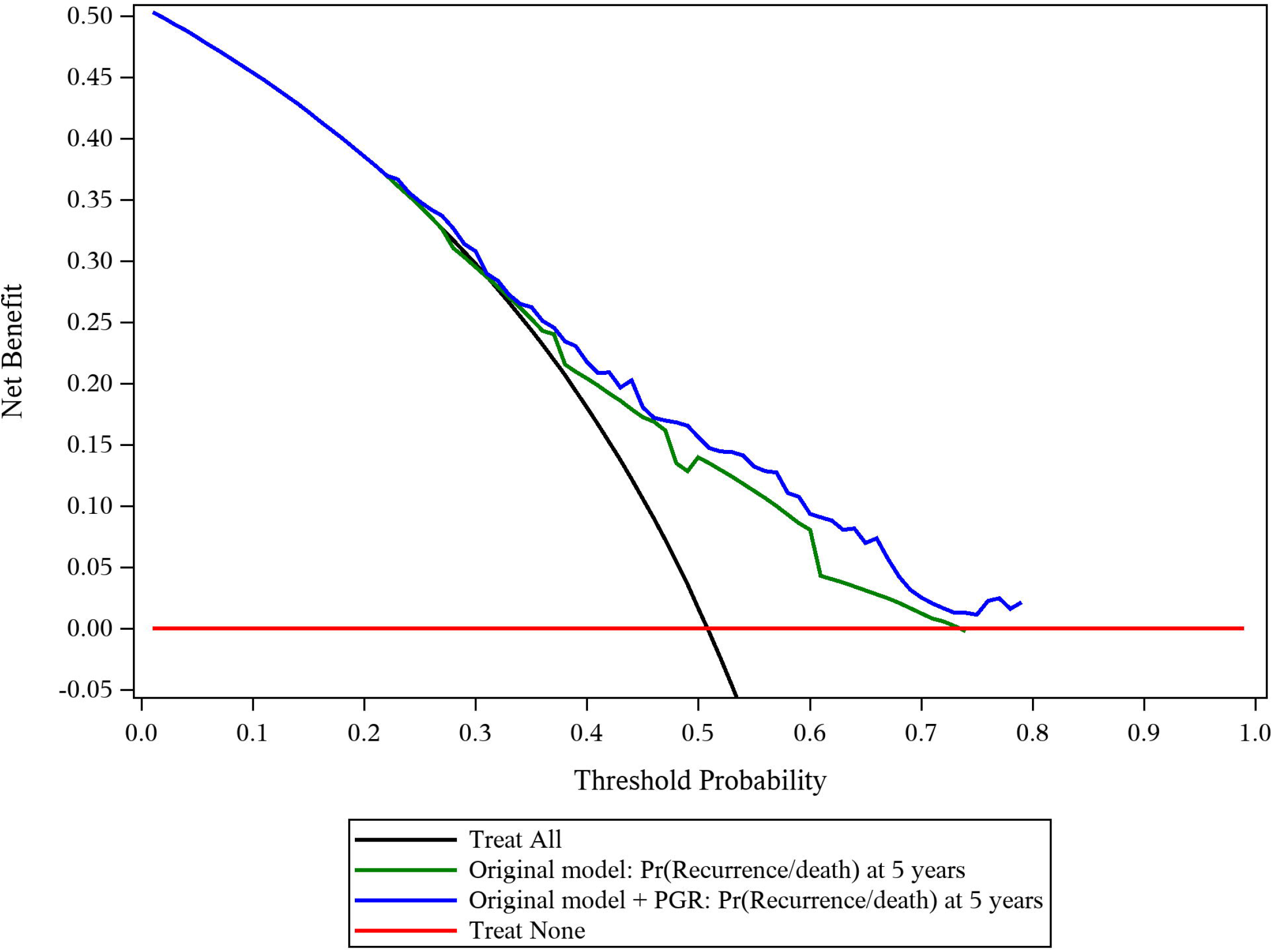
Calibration plot of model predicting recurrence within 5 years for patients with primary breast cancer in external validation data for A) Fixed time assessment. Decision curves for predicted probabilities without (green line) and with (blue line) PGR in B) development dataset; C) external validation dataset. A External validation: Fixed time assessment (predicted risk at 5 years from original model versus secondary model) B Decision curve analysis in development data C Decision curve analysis in external validation data Footnote: In part A, the solid red line represents a restricted cubic spline between the predicted risk from the developed model and the predicted risk from the refitted Cox model at 5 years. The dashed lines represent the 95% confidence limits of the predicted risks from the refitted model. At the bottom of the plots is the density function for the predicted risk from the developed model.

#### Strong calibration

Ideally, we would check for strong calibration by comparing predictions to the observed event rate for every covariate pattern observed in the validation data. However, this is hardly ever possible due to limited sample size and/or the presence of continuous predictors.

#### Time range calibration

Mean calibration can be assessed by comparing observed to predicted event counts, a method that is closely related to the standardized mortality ratio (SMR), common in epidemiology.^28, 29^ For the validation cohort, the total number of observed recurrent free survival endpoints was 285 versus an expected number of 269.9 (ratio 1.06 [0.94 to 1.19]). This agrees with the 5 year fixed time results. For weak and moderate calibration assessment, a similar path to the fixed time approach can be followed using a Poisson model with the predicted cumulative hazard from the original Cox model as an offset.^11^ The weak calibration results gave a calibration slope of 1.05 [95% CI 0.80 to 1.30] respectively, again confirming very good calibration. Computational details are in Appendix 3.

### Overall performance

Another common measure used at validation of predictions up to a fixed time point, encompassing both discrimination and calibration, is the Brier score.^30–32^ This measure also involves inverse weights and is the mean squared difference between observed survival at a fixed time point (event =1 or 0) and the predicted risk by that time point.

The Brier score for a model can range from 0 for a perfect model to 0.25 for a non-informative model in a dataset with a 50% event rate by the fixed time point. When the event rate is lower, the maximum score for a non-informative model is lower, which complicates interpretation. A solution is to scale the Brier score, B, at 0 – 100% by calculating a scaled Brier score as 1-B/B_0_, where B_0_ is the Brier score when using the same estimated risk (the overall Kaplan-Meier estimate) for all patients.^33^

At apparent validation, the Brier score was 0.210 [95% CI 0.204 to 0.216], with a null model Brier score B_0_ of 0.245, so a scaled Brier score of 14.3% [95% CI 11.8% to 16.8%]. The internal validation results were very similar to the apparent validation. At external validation, the Brier score was slightly higher at 0.224 [95% CI 0.210 to 0.240] and the scaled Brier score lower at 10.2% [95% CI 4.0% to 15.9%] (Table 3).

### Approaches to assess clinical usefulness

Measures of discrimination and calibration quantify a model’s predictive ability from a statistical perspective. However, they fall short with regard to evaluating whether the model may actually improve clinical decision making.^34–36^ Specifically, we may wish to determine whether a model is useful to support targeting of an additional treatment to high risk patients. This is what decision curve analysis aims to do by calculating the Net Benefit of a model.^36, 37^ First, we need to define a clinically motivated risk threshold to decide who should be treated. For example, we may offer chemotherapy to patients with a 5-year risk of recurrence or death exceeding 20%. Using this 20% threshold, treatment benefit is obtained for patients who would die or whose cancer would recur within 5-years and have a risk ≥20%: true positive classifications. Harm of unnecessary treatment is caused to those patients who would not die or whose cancer would not recur within 5-years but have a risk ≥20%: false-positive classifications. ^38^If the harm of unnecessary treatment (i.e. a false positive decision) is small then a risk threshold close to 0% is sensible, as it would lead to treating most patients. However, if overtreatment is harmful, such as major surgery, then a higher risk threshold may be apt. The odds of the risk threshold equals the harm-to-benefit ratio. Realizing this, we can now calculate the Net Benefit by calculating the proportion of true positives (that benefit) and substracting from that the proportion of false positives (that are harmed), weighted by the harm-to-benefit ratio (*w*): ^38^

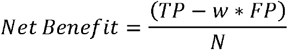

where *TP* is the number of true-positive decisions, *FP* the number of false-positive decisions, *N* is the total number of patients and *w* is the odds of the threshold. When we are dealing with survival data, the Net Benefit can be calculated in the presence of censoring at any prediction horizon (Vickers et al, 2008).^35^ For survival data *TP* and *FP* are calculated as:

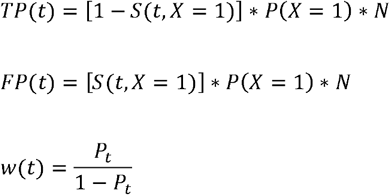

where *P*_*t*_ is the predicted probability at time *t*, 1 – S(t, X=1) the observed event probability for those classified as positive, and P(*X=1)* is the probability of a positive classification.

Considering only one single risk threshold for evaluation of Net Benefit is usually too limited, since the perceived harms and benefits of treatment may differ between decision makers and be context-dependent. Hence, we specify a range of reasonable thresholds which would be acceptable for treatment decisions.^39^ The Net Benefit can be visualised for this range of clinically relevant thresholds using a decision curve. Decision curve analysis allows us to compare the Net Benefit for different prediction models to the default strategies of treating all or no patients (‘treat all’ and ‘treat none’).^37, 40 7^

Based on previous research we focused on a range of thresholds from 14% to 23% for adjuvant chemotherapy (Figure 1B).^41^ If we choose the threshold of 23% the model has a Net Benefit of 0.27. This means that the model would identify 27 patients per 100 who will have recurrent breast cancer or die within 5 years of surgery and thus require adjuvant chemotherapy. The decision curve based on the development data shows that the model Net Benefit is only marginally greater than a ‘treat all’ reference strategy at the highest threshold within the acceptable range of 23%. However, in the external validation dataset, the model is not useful as it has similar Net Benefit values to the ‘treat all’ strategy for the full range of clinically acceptable thresholds. Therefore it is unlikely that the model is useful to support decisions around adjuvant chemotherapy (Fig 1C).

All the methods we have described are summarised in the Appendix (Table S2).

### Model extension with a marker

We recognize that a key interest in contemporary medical research is whether a particular marker (e.g. molecular, genetic, imaging) adds to the performance of an existing prediction model. Validation in an independent dataset is the best way to compare the performance of a model with and without a new marker. We extended our model by adding the progesterone (PGR) biomarker at primary surgery to the Cox model (Table 2). The results are described in appendix 4 and presented in Table 3. Briefly, at external validation the improvement in fixed time point discrimination was from 0.693 to 0.722 (delta AUC of 0.029), the improvement in time range discrimination was from 0.639 to 0.665 (delta C of 0.026). There was an improvement in net benefit (0.367 versus 0.362), which means we need to measure PGR in 200 patients for one additional net true positive classification.

#### Software

All analyses were done in SAS v 9.4 (SAS Institute Inc., Cary, NC, USA) and R version 3.6.3, R Foundation for Statistical Computing, Vienna, Austria). Code is provided at https://github.com/danielegiardiello/Prediction_performance_survival.

## Discussion

This article provides guidance for different measures that may be used to assess the performance of a Cox proportional hazards model. The performance measures were illustrated for use at model development and external validation. At model development, the apparent performance can directly be assessed for a prediction model, and internal validity is commonly assessed by cross-validation or bootstrapping techniques. External validation is considered a stronger test for a model. We first illustrated how to evaluate the quality of predictions using measures of discrimination, calibration and overall performance. We then showed how to evaluate the quality of decisions according to Net Benefit and decision curve analysis. Finally, we illustrated that the performance measures are also applicable when assessing the added value of a new predictor, where specific interest may be in improvement in discrimination and Net Benefit.

We made a distinction between measures that can be used to assess the performance of predictions for specific time points (e.g. 5- or 10-year survival) and over a range of follow up time. Prediction at specific timepoints will often be most relevant since clinicians and patients are usually interested in prognosis within a specified period of time. As described, AUC, smooth calibration curves and Brier score focus on such specific time points. Of note, estimation of the baseline survival is treated as an optional extra step in most statistical software packages. The consequence is that such key information is not available for most prediction models that are based on the Cox model. This may lead to the misconception that the Cox model does not give estimates of absolute risk. If the baseline survival for specific times points is given together with the estimated log hazard ratios, external validation is feasible (see Table S3). The discrimination and Brier score methods presented here can easily be applied to parametric survival models such as Weibull or more flexible approaches^42^

In the breast cancer study, the optimism in all performance measures was minimal at internal validation. This reflects the relatively large sample size in relation to the small number of predictors, which allows for robust statistical modeling. The performance at external validation was slightly poorer, as can in general be expected and may reflect slightly differential prognostic effects, but also differences in case-mix and censoring distribution.^43^ We have not addressed the common problem of missing values for predictors, which needs somewhat more complex handling than for binary outcome prediction.^44^

Dealing with censoring is a key challenge in the assessment of performance of a prediction model for survival outcomes. If censoring is merely by end of study period (‘administrative censoring’), the assumption of censoring being non-informative may be reasonable. This may not be the case for patients who are lost to follow-up, where censoring may depend on predictors in the model and other characteristics. As well as the IPCW and secondary modelling approaches presented here, other approaches are possible, for example using pseudo-observations, which often makes the assumption of fully uninformative censoring. Extensions that can deal with covariate-dependent censoring have been proposed.^45, 46^

### Recommendations

We provide some recommendations for assessing the performance of a survival prediction models (Box 2 and Table S3). For calibration at external validation, we recommend plotting a smooth calibration curve (moderate calibration) and reporting both mean and weak calibration. Where no baseline survival is reported from the development study, only crude visual calibration and discrimination assessment may be possible (Appendix 5). Moreover, we recommend that researchers developing or validating a prognostic model follow the TRIPOD checklist to ensure transparent reporting.^7^

#### Box 2. Recommendations for assessing performance of prediction models for survival outcomes

***Assessment***

- For overall performance, we recommend reporting a scaled Brier score for a fixed time point assessment.
- For discrimination, report time-dependent area under the ROC curve at the time point(s) of primary interest. We recommend Uno’s weighted approach. For assessment over a time range we recommend either Harrell’s C or Uno’s C.
- For calibration in an external dataset, while moderate calibration is essential, we recommend following the calibration hierarchy and also reporting mean and weak calibration.

***Clinical utility***

- If the model is to support clinical decision making, use decision curve analysis to assess the Net Benefit for a range of clinically defendable thresholds.

***Publication***

- When reporting development of a prediction model, include the baseline survival and ideally a link to a dataset containing the full baseline survival so others can validate the model at a fixed time point or over a range of follow up time. Report model coefficients or the hazard ratios. Both baseline survival and coefficients are essential for independent external validation of the model.
- Use the TRIPOD checklist for reporting prediction model development and validation.

Net Benefit, with visualisation in a decision curve, is a simple summary measure to quantify the potential clinical usefulness when a prediction model intends to support clinical decision-making. Discrimination and calibration are important but not sufficient for clinical usefulness. For example, the decision threshold for clinical decisions may be outside the range of predictions provided by a model, even if that model has a high discriminatory ability. Furthermore, poor calibration can ruin Net Benefit, such that using a model can lead to worse decisions than without a model.^47^

We recognize that other performance measures are available that have not been described in this paper, which may be important under specific circumstances. We recommend that future work should focus on assessing performance for various extensions of predicting survival, such as for competing risk and dynamic prediction situations.^22, 48–51^

In conclusion, the provided guidance in this paper may be important for applied researchers to know how to assess, report, and interpret discrimination, calibration and overall performance for survival prediction models. Decision curve analysis and Net Benefit provide valuable additional insight on the usefulness of such models. In line with the TRIPOD recommendations, these measures should be reported if the model is to be used to support clinical decision making.

## Data Availability

All data produced are available within the R package 'survival'

