## Supplementary material for "Assessing performance and clinical usefulness in prediction models with survival outcomes: practical guidance for Cox proportional hazards models"

**Appendix 1: Types of validation**

***Apparent performance***

Apparent performance is the model’s performance estimated on the same data that was used for developing the model. It is usually optimistic and therefore a poor estimate of the predictive performance in new individuals, even if those individuals are from the same population. The ultimate aim of a prediction model is to apply it on new patients for whom the outcome is still unknown. This is why it is important to conduct internal and external validation.

***Internal validation***

After model development it is important that we at least assess performance of the model’s predictions for patients from the same underlying population.^1^ The most well-known method splits the data into a model development part and a model testing part. The model is developed on the first set of data, and its performance is assessed on the second. While simple and transparent, this method is often inefficient^2^: the available data is split into two smaller parts, such that both model development and performance assessment become more uncertain. It is better to develop the model on all available data to maximize development sample size, and to use resampling methods for internal validation. The most common methods are cross-validation and bootstrapping. Cross-validation is a generalization of the split-sample method which involves splitting the data into groups. With splitting by decile, the model is estimated on 90% of the data and tested on the remaining 10%. This is repeated another 9 times, each time using the next 10% for testing. The average performance is calculated over the 10 repetitions. For more stability, such a 10-fold cross-validation procedure can be repeated 10 times (10x10-fold cv).^3^ Alternatively, internal validation can be done using bootstrapping, which provides even more stable estimates of performance (at the price of increased computation time) for the population where the sample originated from. This method involves generating samples from the underlying population by drawing n samples (in the case study we used n=500) with replacement from the original dataset. Each of the n samples are the same size as the original dataset.^3^ The model development process is repeated in each of the bootstrap samples and their performance assessed (bootstrap performance). Each of the models is then applied to the original dataset and test performance assessed. The average difference in the bootstrap and test performance is the ‘optimism’ in performance of the original model. Optimism-corrected performance is estimated as apparent performance minus optimism. It is an estimate of internal validity, reflecting validation for the underlying population where the data originated from.^4, 5^

***External validation***

It is preferable to have prediction models that are transportable to new (external) populations that are ‘plausibly related’ to those used to develop the model.^6–8^ The simplest example involves the application of the model in patients from a different location. Evaluating this type of external transportability is referred to as geographical validation. Of specific interest is the evaluation of the heterogeneity in performance across many locations.^9^ However, because populations at any given location tend to change over time, for example due to changes in patient care, another type of external validation involves the evaluation of a model in more recent patients from the model development location. This is referred to as temporal validation. In addition to geographical and temporal validation, it may also be relevant to determine whether a model performs well for a different type of population than the one it was developed on (domain validation).^10^ For example, does a model that predicts mortality within 5 years from the point of diagnosis of early breast cancer, predict accurately for patients diagnosed with locally advanced breast cancer?.^11^

Externally validating a survival prediction model is problematic if the published article does not report the estimate of the baseline survival function for any follow-up times.

**Appendix 2 Further details on methods for assessing discrimination**

***Time-dependent AUC***

The standard approach of ROC curve analysis considers outcome status for a patient as being binary. However, in the survival setting the result depends on the timepoint of interest since the proportion of events changes over time. Recent research has incorporated this dependency on time into the estimation of sensitivity and specificity (and hence the AUC). This means that since the disease status can be observed at each time point, we may obtain different values of sensitivity and specificity throughout follow-up. This may be useful to determine how well the model performs for patients early in follow-up compared to longer term survivors. Three different approaches to estimating time-dependent sensitivity and specificity have been proposed. Each differ with regards to the time-dependent manner that the outcome status is handled.^12^ In prognostic modelling the goal is generally to predict an outcome that occurs within a time period of clinical interest (e.g. within 5 years in our case study). Under this scenario we propose to focus on one suitable approach to estimate sensitivity and specificity (and hence the AUC) called ‘cumulative sensitivity and dynamic specificity’. Here, at each time point each patient is classed as either a case or a non-case where a case is a patient who experiences the outcome between baseline and the time point of interest, t (e.g., 5 years), and a non-case is a patient who remains outcome free at t. The AUC evaluates whether predicted probabilities were higher for those who experience the outcome at or prior to t than for those who still have to experience the outcome.^12^

The Kamarudin review identified eight methods of evaluating the time-dependent AUC using the cumulative sensitivity and dynamic specificity approach and we illustrate one in our case study that is recommended by Blanche et al, 2013;^12, 13^ the inverse probability of censoring weighting approach by Uno et al, 2007.^14^ This approach allows us to reassign the case weights of those censored to other observations with longer follow up (see Table S1 for details of various methods for dealing with censored patients).

***Concordance***

Concordance (C) is one of the most popular measures of discrimination. C is defined as the fraction of all pairs of observations for which the rank order of the predictions agrees with the rank order of the actual response, i.e., the prediction model got them in the right order. Observation pairs that have the same response are not used, while pairs that have the same predicted value count as 1/2 an agreement. For a continuous response this definition is equivalent to Somers' d, for a binomial response it leads to the area under the curve (AUC), and for a survival response to Harrell's C. C is only equivalent to the AUC for binomial outcomes which has caused confusion for applied researchers who incorrectly use these terms interchangeably in the survival setting.^15^ For survival data, Harrell’s C is the most commonly applied, however, it does not account for censored data. Two important refinements to C for survival data are the addition of administrative censoring at the time point of interest, *t*, and the addition of a time dependent weighting that more fully adjusts for censoring.^16^ If interest is focused on predicted survival up to *t*=5 years, for instance, then relative rankings between patient pairs who both have events beyond 5 years might be considered irrelevant. For the example data, the estimated 5-year concordance for prediction in the development and validation data sets was 0.674 (95% CI 0.660 to 0.688) and 0.652 (95% CI 0.619 to 0.685), respectively, using Harrell’s C]). Uno’s C uses a time dependent weighting that more fully adjusts for censoring. Using Uno’s C, the estimated 5-year concordance was 0.673 (95% CI 0.657 to 0.689) in the development data and 0.639 (95% CI 0.602 to 0.676) in the external data. It has been shown that the bias from Harrell’s C is more pronounced when it is greater than 0.8 which is rare for prediction modelling in the absence of overfitting.^17^ Weighted measures such as Uno have been shown to become biased when censoring is large leading to extreme weights.^17^

**Table S1 Approaches to deal with censoring in the analysis of performance at a fixed time point for a survival outcome**

| **Approach** | **Concept** | **Assumption** | **Applications** | **Data illustration ^** |
| --- | --- | --- | --- | --- |
| Inverse probability of censoring weights (IPCW) | Set the weights of patients censored before time *t* to zero, reassigning their mass to other patients still at risk at time *t.*  Can also be extended to a time dependent IPCW. | Fully uninformative censoring* | Weighted Brier score; Uno’s approach to discrimination  Uno’s C uses a time dependent weighting (more details in appendix 2)^16^ | Redistribute the weight of 280 patients who are censored before 5 years to the 406 with either an event or no event observed at 5 years |
| Use of a secondary model | Impute censored observations by predictions from a flexible secondary model using the complementary log-log transformed predicted risk at t years as the only covariate. | Uninformative censoring given the risk score, and proportional hazards ** | Austin et al (2020) approach to calibration.^18^ | Analyze 686 patients |
| Pseudo values | Impute censored patients by estimated survival captured in pseudo values | Fully uninformative censoring but extensions can deal with covariate-dependent censoring. | Assess calibration and discrimination with pseudo values | Analyze 686 patients (including 280 censored patients) with pseudo values |

^ 280/686 GBSG (external validation dataset) subjects are censored before 5 years

* This assumption is stronger than at model development, where censoring is assumed to be uninformative given the risk score (as modeled from predictors or outcome). However, methods are available to make the weights covariate dependent^19^

** This assumption is similar to model development with Cox regression.

**Table S2 Characteristics of key performance measures for the evaluation of survival prediction models**

| ***Aspect*** | ***Fixed time point or time range*** | ***Measure*** | ***Visualization*** | ***Characteristics*** |
| --- | --- | --- | --- | --- |
| Discrimination | Fixed | Time-dependent (cumulative/dynamic) AUC   - Uno* | Time-dependent AUC curve plots | At time, t, each patient is classed as either a *case* or a *non-case*. A case is a patient who experiences the outcome between baseline and t (or at t). A non-case is a patient who remains outcome free at t. The AUC evaluates whether predicted probabilities were higher for those who experience the outcome at or prior to t than for those who still have to experience the outcome.^14, 17, 20^ |
|  | Time range | Concordance (C)   - Uno - Harrell | Kaplan Meier curves provide informal evidence of discrimination^21^ (Appendix 6) | Calculated as a fraction where the denominator is the number of all possible pairs of patients in which one patient experiences the event first and the other later. C quantifies the degree of concordance as the proportion of such pairs where the patient with a longer survival time has better predicted survival.^20^ Harrel’s C excludes pairs where the patient with shorter follow up is censored. Uno’ C adjusts more fully for censoring.^16^ |
| Calibration | Fixed  Time range | *Mean calibration (calibration-in-the-large)*   - (1-Kaplan-Meier)/average predicted risk at t - Poisson model intercept (O/E) |  | Simplest type of calibration which evaluates if the observed outcome rate is equal to the average predicted risk.  Use Poisson model intercept with log cumulative hazard as offset.^22^ |
|  | Fixed  Time range | *Weak calibration*   - Calibration slope using secondary Cox model - Calibration slope using Poisson model |  | Assesses global under or over prediction and overfitting (slope<1) or underfitting (slope>1). See appendix 3 for details on calculations.  Slope is coefficient of PI in Poisson model with log cumulative hazard function minus PI as offset. |
|  | Fixed  Time range | *Moderate calibration*   - Model relationship between predictions and observed risk in external dataset using secondary Cox model - Complemented with ICI, E50, E90 - Plot of time versus O/E - Model relationship between predictions and observed risk in external dataset using Poisson model | Smooth calibration curve of observed t-year risk of the outcome versus predicted probability by t-years.  Plot the observed / expected number of events over time.  Plot cumulative hazard from Poisson model versus cumulative hazard from original Cox model | Reveals miscalibration which cannot be detected using calibration-in-the-large and the calibration slope approaches. Plot predicted risk of this model against predicted risk from original model.^18^  Visualises O/E across all time points up to t. |
| Overall performance | Fixed | Brier score and scaled Brier score |  | Captures calibration and discrimination aspects.  Interpretability is improved by scaling between 0 and 100%. |
| Clinical usefulness | Fixed | Net Benefit | Decision curve | Net number of true positives gained by using model compared to no model at a single threshold (NB) or over a range of thresholds (DCA)^23^ |

^§^ PI = prognostic index; * A modified version of Uno’s weighted approach is available that uses weights that are the conditional probability of being uncensored. These are calculated using the Cox model and allowing for covariate-dependent (as opposed to uninformative) censoring.^13^

**Appendix 3 Calibration assessment**

Calibration can be evaluated either across all follow up time points (time range assessment) or at one specific time point. Time range assessment refers to the evaluation of estimated risks at the time of the event (or censoring) for each patient. Evaluating models over the time range requires the availability of the development dataset, or at least the baseline survival for all time points. Here we describe the methods for assessment of calibration over the time range:

***Mean calibration***

When we wish to assess calibration across all time points then one method to deal with this is to consider a comparison of the total number of observed events (O) as compared to expected events (E), counts instead of probabilities. The expected count for each subject is defined as the predicted cumulative hazard for that subject, under the model, up until that subject's event time or censoring. This approach has a long history in epidemiology where sum(observed)/sum(expected) is known as a standardized incidence ratio (SIR).^24, 25^ Such data can be analysed using standard Poisson methods and software (Berry, 1983).^26^ However, in order to estimate the cumulative hazard, the dataset used to develop the original model or, at least, the baseline survival for all time points is required.^22^ Failing that, linear interpolation may be used if the baseline survival is available at several time points.

For the Rotterdam dataset, there are 1275 events within 5 years of study entry. Using the German Breast Cancer Study Group (GBCSG) validation dataset, there are 285 observed events while the Rotterdam model applied to that data predicts 269.9, giving an O/E ratio of 1.06. Using the individual observed (as outcome) and expected values (log cumulative hazard as an offset term) a Poisson model, estimates an intercept term of 0.054 with a standard error of 0.059. The exponential of this value leads to exactly the same O/E estimate of 1.06, and a confidence interval of (0.94, 1.19). Fig S1A shows how O/E changes over time, remaining stable from 18 months.

**Fig S1A Time range assessment of O/E in external dataset**

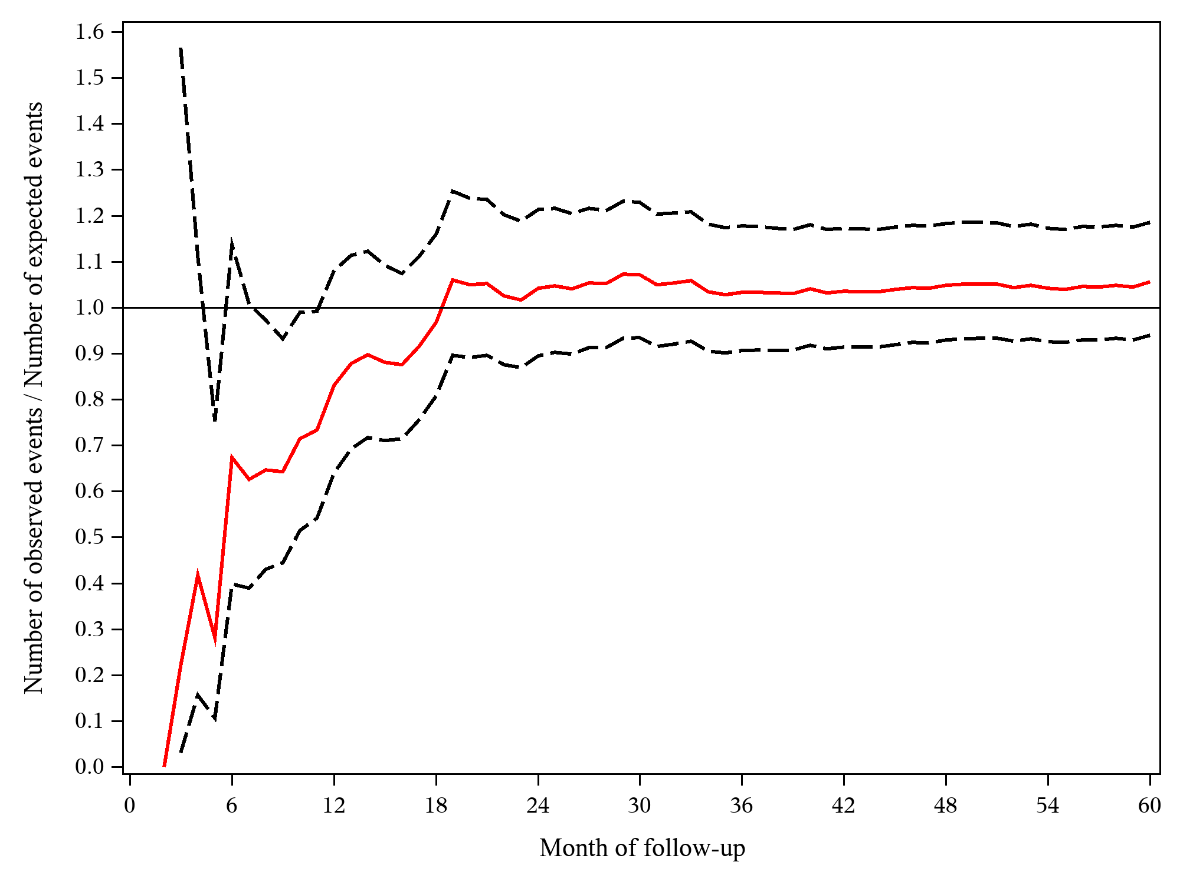

*Note: the solid red line represents O/E at each month up to 5 years and the dashed lines represent the 95% confidence limits of O/E*

***Weak calibration***

For binary outcomes, calibration can be inspected visually using a calibration plot of the observed proportion of outcome associated with a model’s predicted risk. The PI is regressed on the observed outcomes using a logistic calibration model.^27^ The coefficient of PI is the calibration slope and its value indicates whether there is overfitting (slope<1) or underfitting (slope>1).^28^ Since the calibration slope does not involve grouping patients and provides a measure of the magnitude and direction of miscalibration with 95% confidence interval, it is preferred to the Hosmer-Lemeshow goodness-of-fit test, the use of which is discouraged due to focus on p-values and poor test performance characteristics.^28, 29^ For the Cox model, there are different variations of the Hosmer-Lemeshow test including tests proposed by Grønnesby and Borgan,^30^ which should not be used for similar reasons.

For survival outcomes, estimation of the calibration slope is possible using a Poisson model. This is done by including the PI in the validation dataset (using the coefficients from the original Cox model) as a predictor in a Poisson model with the difference between the log cumulative hazard and PI as an offset and using a log link.^22^ The regression coefficient for PI represents the calibration slope. In our study the calibration slope was 1.05 (95% CI 0.80 to 1.30), so close to the ideal value of 1. The calibration intercept is just the intercept term before exponentiating in the previous section on mean calibration. This approach is termed weak calibration because of its limited flexibility in assessing calibration. We are essentially summarising calibration (of the observed proportions of outcomes versus predicted probabilities) using only two parameters. However, more subtle violations of miscalibration may remain undetected.

***Moderate calibration***

The relation between the outcome over the time range and predictions can be visualised by plotting the predicted cumulative hazard from the Poisson model against the predicted risk from the development model. In the external dataset, the PI from the original Cox model is modelled as a restricted cubic spline in a Poisson model with the log of the cumulative hazard as the offset. Predictions from this Poisson model represent a proxy to the observed outcomes for all patients including those who were censored. The calibration plot shows good agreement between the Cox and Poisson models (Fig S1B).

**Fig S1B Calibration plot of predicted cumulative hazard of recurrence over the time range for Cox model versus Poisson model**

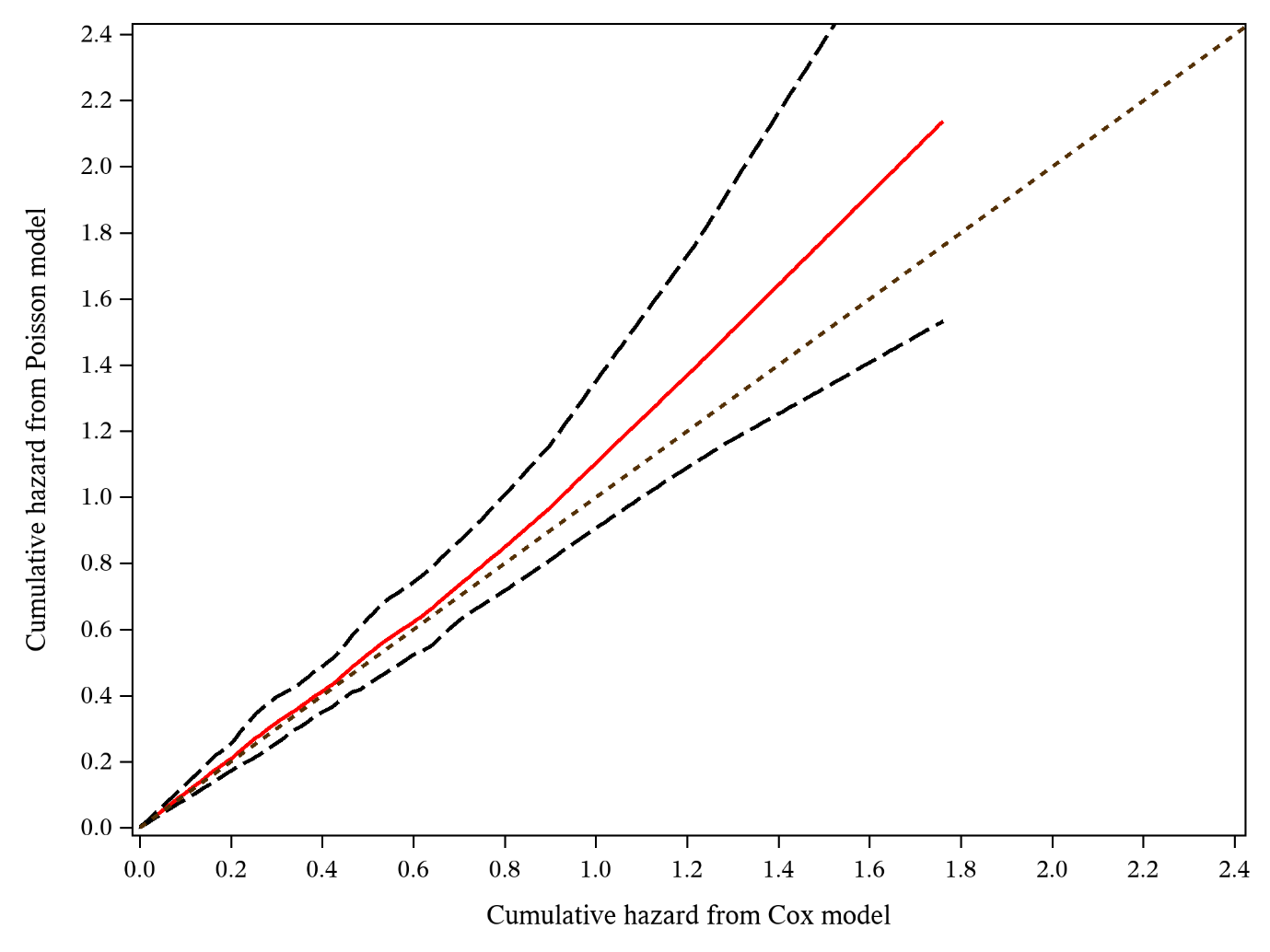

*Note: the solid red line represents the relationship between the predicted cumulative hazard from the developed model and the predicted cumulative hazard from the Poisson model. The dashed lines represent the 95% confidence limits of the predicted cumulative hazard from the Poisson model.*

**Appendix 4 Incremental value of PGR**

We extended the model by adding the progesterone (PGR) biomarker at primary surgery to the Cox model. Following examination for non-linearity, PGR was fitted as a restricted cubic spline function with 3 knots (see Fig S2). We repeated the apparent, internal and external validation processes on this extended model.

***Performance in development dataset***

PGR had additional predictive value when added to the original model, increasing the model chi-squared from 483.7 to 516.7 (LR statistic 33.0, df=2, P<0.001) in the development dataset. Overall performance showed a small increase: Brier score decreased from 0.210 to 0.209, and the scaled Brier score increased from 14.3% to 14.9% (Table 4). The discriminative ability at 5 years follow-up also increased marginally (e.g., Uno’s weight approach increased from 0.712 to 0.720).

For a threshold of 23%, the model with PGR included had a slightly larger net benefit than the model without PGR (0.274 versus 0.267) (Fig 1B). Hence, at this particular cut-off, the model with PGR would be expected to lead to one more net true positive classification per 154 patients (1/0.0065) at the same number of false positive classifications.

***Performance in external dataset***

Comparing the above performance measures for the model with and without PGR in the external dataset, the former was better overall. The improvement in fixed time point discrimination was from 0.693 to 0.722 (delta AUC of 0.029) at external validation while improvement across the time range was from 0.639 to 0.665 (delta C of 0.026). Globally, the total number of observed recurrent free survival endpoints was 285 versus an expected number of 279.0. Using the Poisson model this equated to a calibration-in-the-large SIR of 1.02 (95% CI 0.91 to 1.15)). The calibration slope was 1.16 (95% CI 0.93 to 1.40). Mean calibration on average showed some improvement with PGR included. The calibration plot of O/E across all time points up to 5 years shows relatively consistent results from 18 months onwards (Fig S3A). The calibration plot of the predicted cumulative hazard in the original Cox model versus the Poisson model shows good agreement, although some underprediction in the higher risk patients (Fig S3B). Focusing on calibration at the fixed time point of 5 years we found that the Kaplan-Meier estimate of experiencing the event within 5 years was 0.49, while the average predicted probability was 0.50. The calibration plot (Fig S3C) shows evidence of good agreement overall for predictions of mortality over 5 years. The ICI decreased from 0.03 to 0.02 when PGR was included and E50 dropped from 0.03 to 0.01. The scaled Brier score increased from 10.2% to 13.6% at external validation. Hence a substantial improvement in statistical performance was found.

With PGR in the model, the risk groups are well separated in both the development and validation datasets which implies that the model discriminates well in these cohorts (Figure S4). However, from approximately 3 years into follow-up the middle two risk groups converge for the external dataset.

In the external dataset, the net benefit was similar for models with or without PGR (Figure 1C). However, at the risk threshold of 23% the model without PGR was no better than treating all patients. The model with PGR had a slightly larger net benefit (0.367 versus 0.362), or one additional net true positive classification per 200 patients (1/0.005) at the same number of false positive classifications.

**Fig S2. Plot showing unadjusted (univariable) relations between PGR and predicted probability of recurrence (solid curve) with 95% confidence bands.** The relation was non-linear characterised by a restricted cubic spline function with 3 knots.

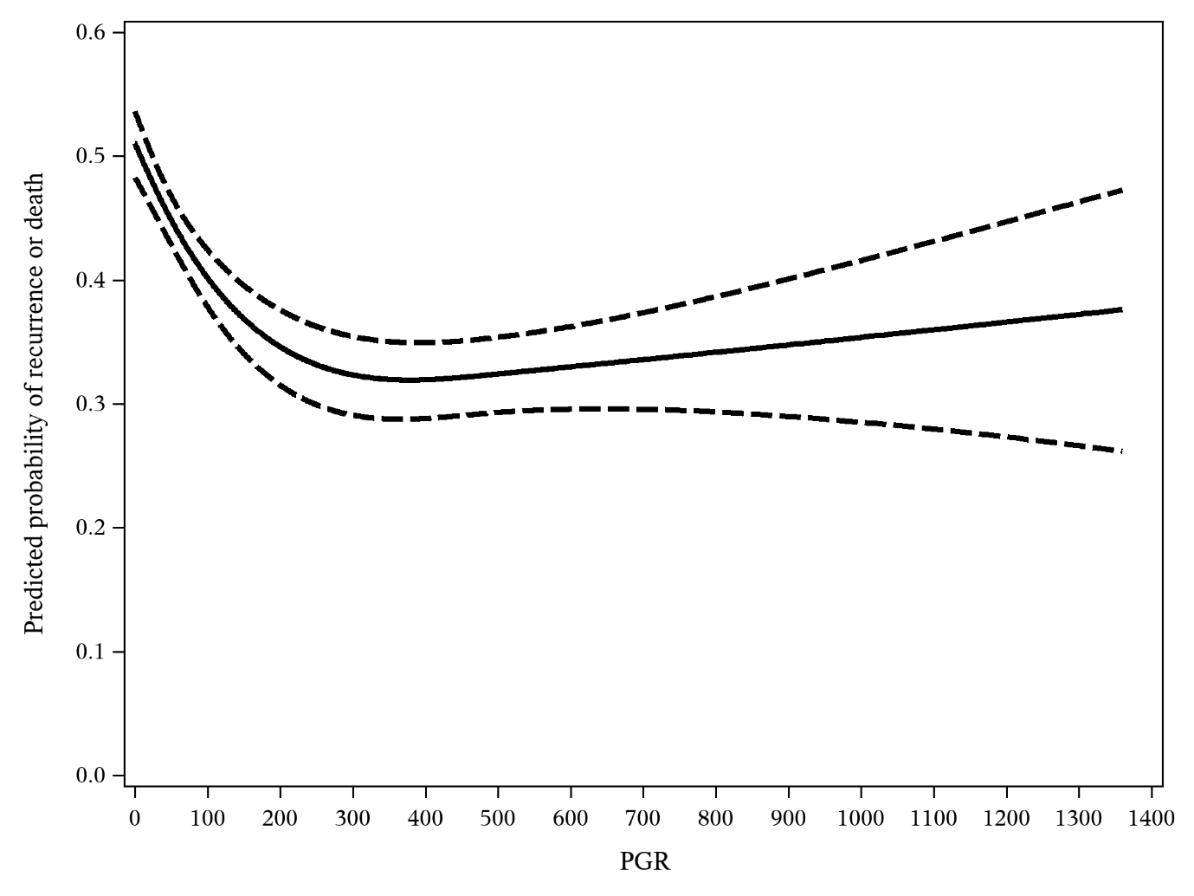

**Fig S3 Calibration plots of Cox model with PGR predicting recurrence within 5 years for patients with primary breast cancer in the external validation data.**

**A O/E across the time range**

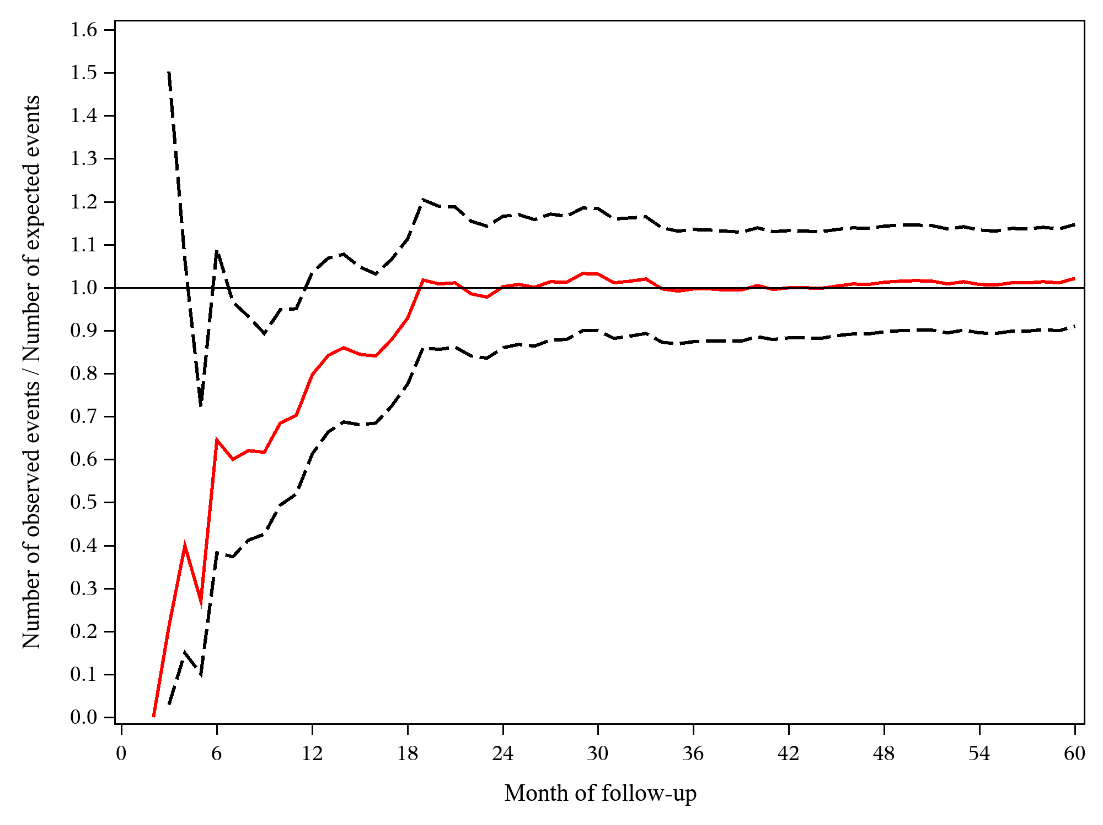

**B Predicted cumulative hazard from original model versus Poisson model**

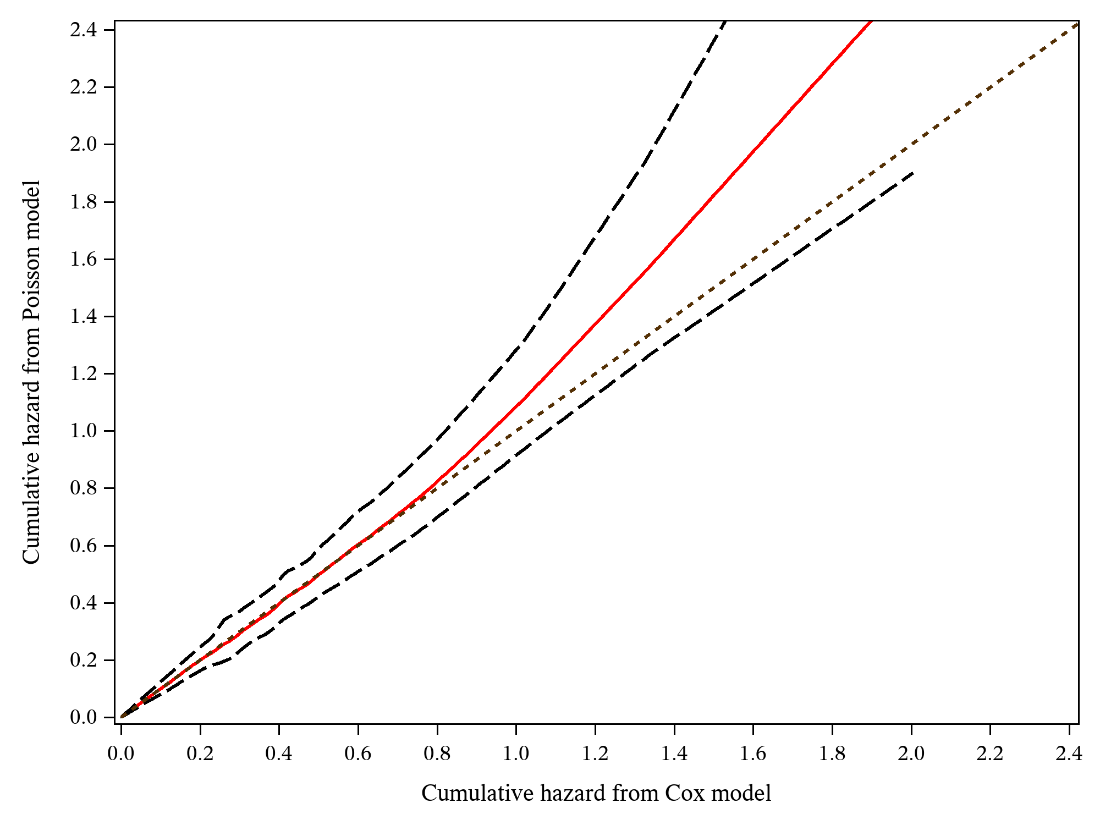

**C Predicted risk from original model versus secondary model at 5 years**

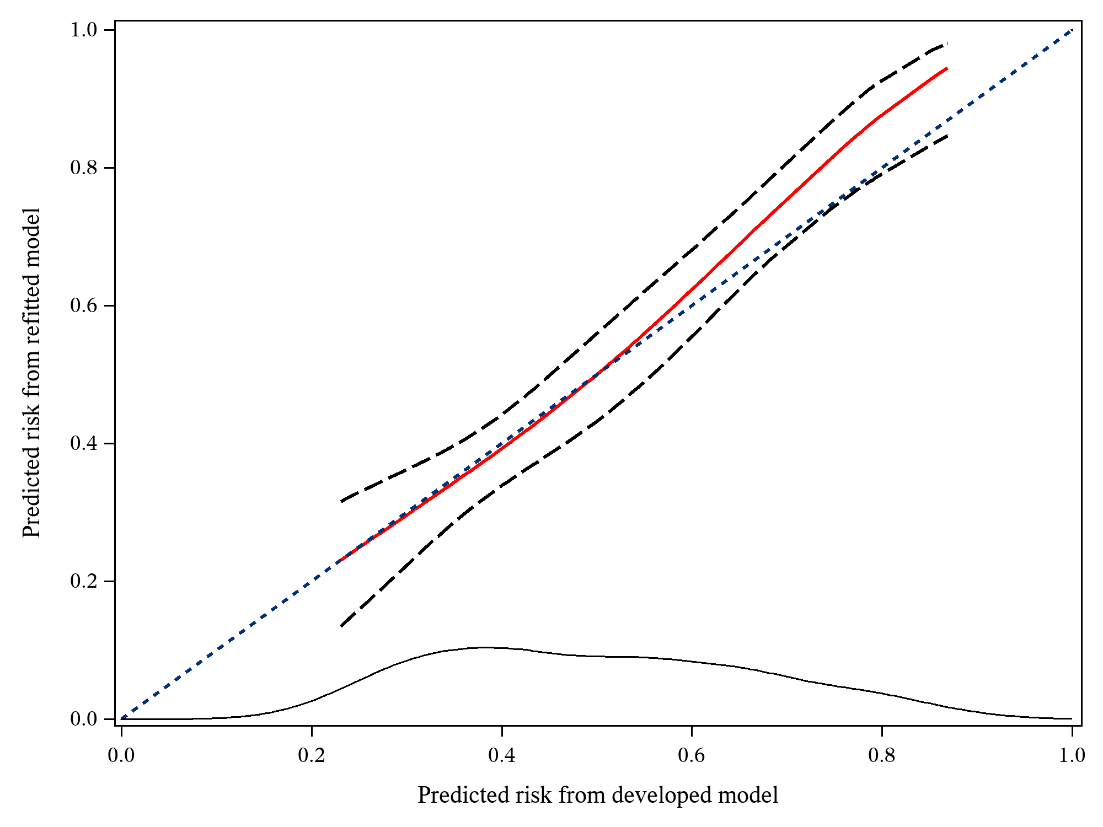

*Note: In A, the solid red line represents O/E at each month up to 5 years and the dashed lines represent the 95% confidence limits of O/E; In B, the solid red line represents the relationship between the predicted cumulative hazard from the developed model and the predicted cumulative hazard from the Poisson model. The dashed lines represent the 95% confidence limits of the predicted cumulative hazard from the Poisson model. In C, the solid red line represents a restricted cubic spline between the predicted risk from the developed model and the predicted risk from the refitted model at 5 years. The dashed lines represent the 95% confidence limits of the predicted risks from the refitted model. At the bottom of the plots is the density function for the predicted risk from the developed model.*

| **What development data do you have?** | **Fixed timepoint assessment** | **Continuous time assessment** | **Methods** |
| --- | --- | --- | --- |
| Whole dataset used to develop model | ***🗸*** | **🗸** | See section on calibration and appendix 3 for calibration methods |
| Table of baseline survival at all observed time points + PI | ***🗸*** | ***🗸*** | See section on calibration and appendix 3 for calibration methods |
| Baseline survival at multiple (but not all) time points (e.g., yearly) + PI | ***🗸*** | ***🗸*** | Use interpolation methods to estimate baseline survival (Crowson et al, 2016).^22^ Then see section on calibration and appendix 3 for calibration methods. |
| A predicted survival curve based on the model + PI | ***🗸*** | ***🗸*** | Use digitisation software to estimate baseline survival (Guyot et al, 2012).^31^ Then see the section of calibration and appendix 3 for calibration methods. |
| Baseline survival at time point of interest + PI | ***🗸*** |  | See section on calibration and appendix 3 for calibration methods at fixed time points. |
| Published Kaplan-Meier curves for risk groups |  |  | Formal assessment not possible. Can visually compare Kaplan-Meier curves to those from validation data (Appendix 5; Royston and Altman, 2013)^21^ |
| None of the above |  |  | Calibration assessment not possible |

**Table S3 What calibration assessments can I do based on the model development information I have?**

**Appendix 5 What to do if the development dataset (or its baseline hazard) is not available**

In case the baseline hazard/survival function (either as a look-up table or mathematical function) of a survival model is not available then there is not enough information to formally assess calibration. However, if the development paper reported Kaplan-Meier curves for risk groups of the PI then it is possible to compare these with the corresponding Kaplan-Meier curves from the validation cohort.^21, 32^ This is not a strict comparison between observed and predicted values since we are using Kaplan-Meier estimates and not the Cox model-based predictions. If the survival curves for risk groups overlap between the development and validation datasets, then this may provide an indication of agreement. Further, plots where the curves are widely separated between risk groups provides informal evidence of discrimination.

In the case study, we centred the PI for the model including PGR at average risk by subtracting its mean of 0.65 and then categorised it into quarters. The groups at the extreme ends represent the lower and upper fourth of the risk of recurrence. This procedure was done in both the development and validation datasets. For the validation dataset the PI was calculated based on the coefficients from the model fitted to the development dataset (Figure S4). In the development dataset the four risk groups are well separated which implies that the model has discriminative ability in this cohort. However, the curves for the second and third fourths are close together in the validation data suggesting that the model does not discriminate well between these two groups. Otherwise, the discrimination is broadly similar between the two datasets. The curves do not agree too well in absolute risks between the two datasets suggesting that there is a degree of miscalibration. The percentage of patients within the four groups in the validation dataset were 8.8%, 21.0%, 36.3% and 34.0% respectively so there are more in the two highest risk fourths and fewer in the lowest risk fourth than in the development dataset. The mean (SD) PI was 0.24 (0.50) in the validation dataset, implying that the prognostic profile was somewhat worse than in the development dataset. This is evident from Table 1 which shows that women in the validation dataset had larger tumours and more nodes.

**Fig S4 Kaplan-Meier curves for event-free survival in 4 equal sized risk groups in the development and validation cohorts for model with PGR**

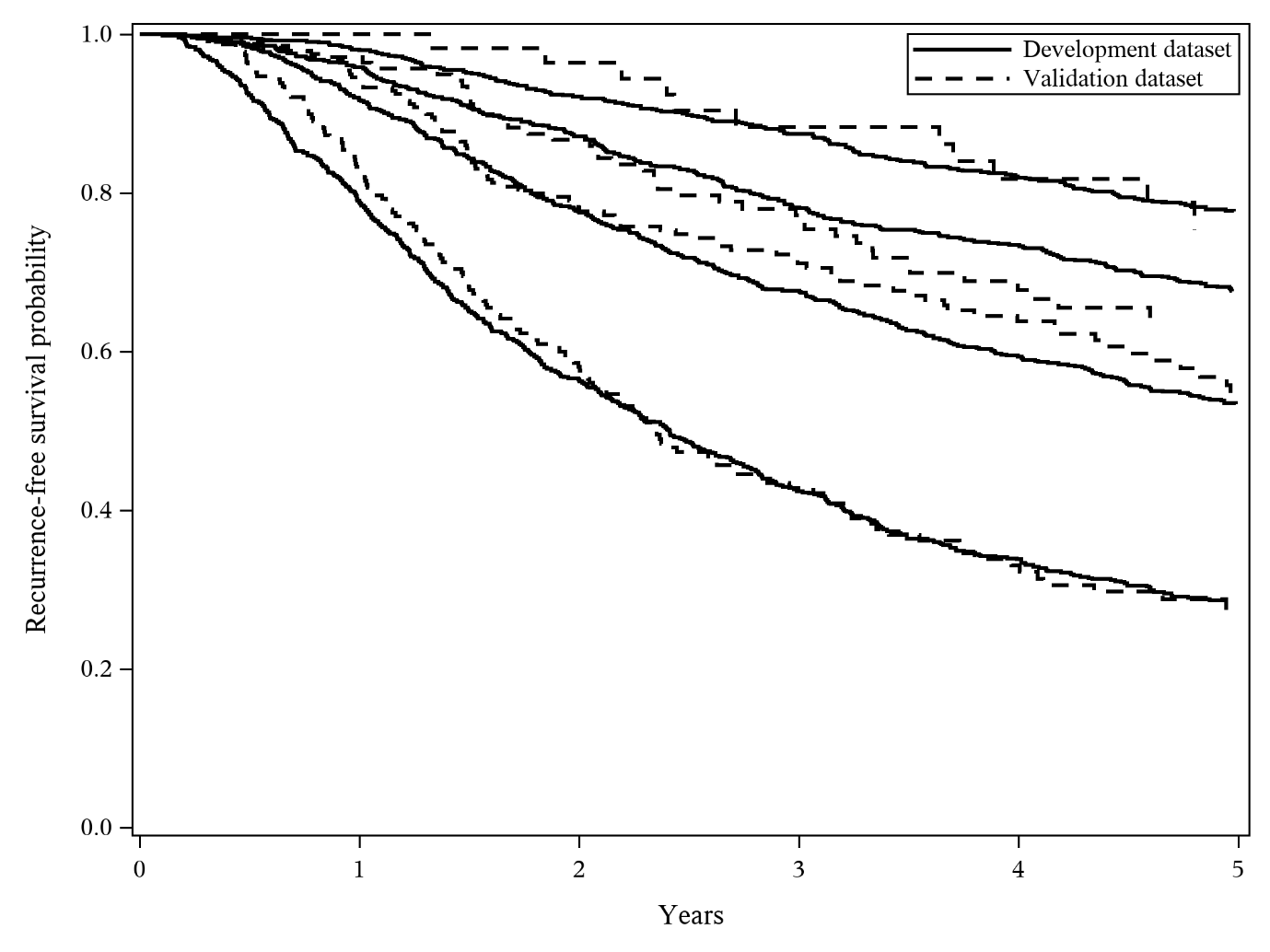
